# Pediatric nicotine exposures from devices and liquids: a comparative analysis of U.S. poison center data

**DOI:** 10.64898/2026.07.04.26357293

**Authors:** Robert S. Miller, Shawn M. Varney

## Abstract

**Introduction:** Pediatric nicotine exposures remain an important and preventable public health issue, particularly with the rapid expansion of electronic nicotine delivery systems. This study compared demographic characteristics, exposure circumstances, and clinical outcomes between pediatric cases involving nicotine devices and bottled liquids reported to U.S. poison centers.

**Method:** This retrospective cohort study analyzed National Poison Data System cases from 2011–2022 involving children aged ≤ 5 years exposed to nicotine devices or bottled liquids. Analyses were limited to cases with definitive medical outcomes. The primary outcome was defined as a moderate or major clinical effect or death. Odds ratios with 95% confidence intervals were calculated, with a secondary analysis restricted to route-concordant exposures.

**Results:** The final cohort included 15,497 cases: 10,168 device exposures and 5,329 liquid exposures. Demographic characteristics were similar between groups. Device exposures more frequently involved inhalation, while ingestion predominated overall. Clinical effects were typically mild and transient, with vomiting and coughing most commonly reported. The primary outcome occurred in 1.9% of device cases and 2.0% of liquid cases (OR = 1.05; 95% CI 0.82–1.34). A secondary analysis restricted to inhalation-only device exposures and ingestion-only liquid exposures similarly found no significant difference in clinically important outcomes (OR = 1.38; 95% CI 0.92–2.12). Two deaths occurred, one in each group.

**Conclusion:** These findings suggest that, despite differences in formulation and route of exposure, nicotine devices and bottled liquids produce broadly similar clinical toxicity profiles in young children. Prevention strategies should address all household nicotine products rather than focusing on specific delivery systems.

## INTRODUCTION

Nicotine is a well-established toxicant, and young children are at significant risk for unintentional exposure due to their natural exploratory behavior [1,2]. While classic pediatric exposures involved traditional tobacco products, the modern landscape has evolved dramatically, driven by a growing consumer market and the development of nicotine salt formulations that allow for substantially higher concentrations and potency [3–5]. Consequently, these novel products present a unique challenge, as historical clinical management recommendations largely predate the widespread availability of high-concentration nicotine salt formulations [6,7]. The toxic effects are mediated through a biphasic action at nicotinic acetylcholine receptors, which can rapidly progress from nausea and tachycardia to severe effects like seizures and respiratory depression [8,9].

Over the past two decades, the popularity of electronic nicotine delivery systems (e-cigarettes) has surged globally, evolving from early “cig-a-like” models to powerful, modern devices that take advantage of substantial improvements in battery technology [10,11]. This trend has fundamentally altered pediatric exposures, shifting the primary risk from ingestion of bottled e-liquids to include direct inhalation from devices [12]. These different exposure pathways raise competing toxicological concerns. On one hand, pulmonary absorption bypasses first-pass metabolism and produces rapid systemic absorption [8]. Conversely, inhalation may be self-limiting in a child, while the large volumes and high concentrations of e-liquids pose a significant risk following ingestion.

The epidemiological studies by Olivas et al. [2] and Govindarajan et al. [3] have provided timely updates on pediatric nicotine exposures by characterizing national trends, identifying high-risk populations, and documenting the evolving contribution of electronic nicotine delivery systems. In these investigations, nicotine devices and bottled e-liquids were grouped because the primary objective was to describe the overall burden of pediatric nicotine exposures rather than to distinguish risks associated with specific exposure pathways. We agree that this approach is appropriate for most epidemiological applications. However, whether these different exposure pathways produce clinically meaningful differences in toxicity remains uncertain, limiting their usefulness for poison center triage and disposition decisions.

Given these differences, we hypothesized that unintentional exposures involving devices would be associated with lower clinical severity than exposures involving bottled e-liquids. This hypothesis reflects the expectation that inhalational exposures are limited in duration and dose relative to liquid ingestion. To address this question, we compared the clinical severity of pediatric nicotine exposures involving e-cigarette devices versus bottled e-liquids. We also performed a secondary, route-restricted analysis comparing inhalation-only device exposures with ingestion-only liquid exposures to better isolate the toxicological effects of the predominant exposure pathways and generate findings more directly applicable to poison center triage and referral decisions.

## METHODS

### Data source and study design

This retrospective cohort study analyzed pediatric nicotine exposures reported to the National Poison Data System (NPDS) between January 1, 2011, and December 31, 2022. NPDS is a comprehensive, near real-time database that aggregates exposure records from all U.S. poison centers. Cases were eligible for inclusion if the exposure occurred in a child ≤ 5 years old, involved an acute ingestion or inhalation/nasal exposure, and was assigned a nicotine-related American Association of Poison Control Centers (AAPCC) generic product code. Eligible codes included 0310094, 0310031, and 0200620 for nicotine-containing devices and 0310095, 0310032, and 0200622 for nicotine liquids. Only cases with a definitive medical outcome (not lost to follow-up or with an unknown outcome) were retained as the followed cohort, which served as the analytic sample for all primary and secondary comparisons.

This study protocol was reviewed by the UT Health San Antonio Institutional Review Board (STUDY00002603). It was granted an exemption from formal review on the basis that it exclusively analyzed non-identifiable, de-identified data. This study was conducted and reported in accordance with the Strengthening the Reporting of Observational Studies in Epidemiology (STROBE) guidelines where applicable. Because NPDS is a structured surveillance database derived from coded poison center case records rather than individual chart abstraction, some STROBE elements related to exposure measurement and outcome verification were not directly applicable.

Demographic variables included age, weight, and gender. Patient age was standardized to months and weight to kilograms. Exposure records were then classified into two exposure arms: device (handheld nicotine delivery systems containing a chamber, heating element, and mouthpiece) or liquid (bottled nicotine solutions of various concentrations) based on NPDS generic product codes. Cases lost to follow-up or with an unknown medical outcome were excluded from inferential analyses. Exposure-related variables included exposure site, caller site, management site, and route of exposure (ingestion or inhalation/nasal), which were not mutually exclusive. Clinical effects and therapies were treated as binary indicators, with any occurrence coded as present for that case. Duration of clinical effects was dichotomized as ≤ 8 hours or > 8 hours. Cases with unknown duration were excluded from duration-specific analyses. Primary comparisons were stratified by exposure type (device versus liquid) and performed on the followed cohort.

The primary outcome was defined as the occurrence of a moderate clinical effect, major clinical effect, or death. As a secondary analysis, a route-restricted cohort was constructed to better isolate the predominant exposure pathways. This cohort included only inhalation-only exposures involving nicotine devices and ingestion-only exposures involving nicotine liquids, while excluding device ingestions, liquid inhalations, and cases coded with both ingestion and inhalation/nasal routes.

### Statistical analysis

Continuous variables were summarized as mean and standard deviation and compared between device and liquid exposures using the Wilcoxon rank-sum test; rank-biserial correlation with 95% confidence intervals was reported as the effect size. Categorical variables were summarized as counts and percentages and compared using Pearson’s chi-square test. Fisher’s exact test was used for sparse 2×2 tables, and chi-square tests with Monte Carlo simulation were used for larger tables with small expected counts. Cramér’s V with 95% confidence intervals was reported as the effect size. Clinical effects and therapies were screened using unadjusted two-sided tests; p-values were not corrected for multiple comparisons because these analyses were exploratory. The primary outcome, defined as a moderate clinical effect, major clinical effect, or death, was compared between exposure groups using Fisher’s exact test, with odds ratios and 95% confidence intervals reported. The same analytical approach was applied to the route-restricted secondary cohort. Statistical significance was defined as p < 0.05. All analyses were performed in R (version 4.3.1; R Core Team, 2023) using the RStudio integrated development environment (Posit Team, 2023).

## RESULTS

A total of 26,228 pediatric nicotine exposure cases were initially identified from the NPDS. After applying exclusion criteria, including non-acute exposures and routes other than ingestion or inhalation, 815 cases were removed, yielding 25,413 cases for analysis (figure 1). Of these, 15,497 were classified as followed cases, defined by the availability of a definitive medical outcome. The final analytic cohort was stratified by exposure to either nicotine devices (n = 10,168) or nicotine liquids (n = 5,329). Total exposures increased substantially over the study period, particularly due to a rise in device-related exposures beginning around 2018 (figure 2). Liquid exposures remained relatively stable throughout.

**Figure 1.**
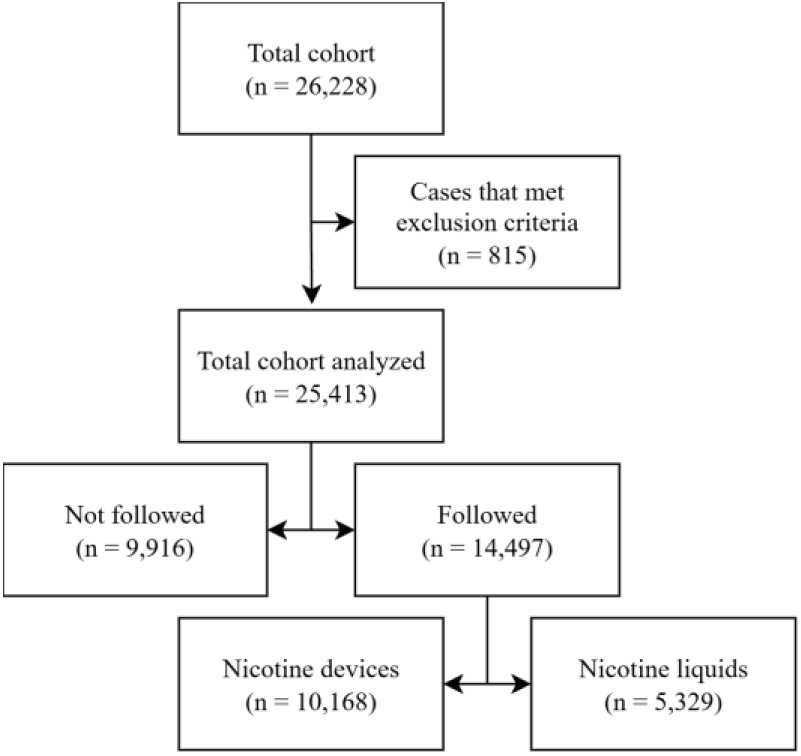
A total of 26,228 pediatric nicotine exposure cases were initially identified. After excluding 815 cases that did not meet inclusion criteria (e.g., non-acute chronicity, exposure routes other than ingestion or inhalation), 25,413 cases remained for analysis. The primary analysis was restricted to the 15,497 cases that were “Followed,” defined as having a definitive medical outcome. This final cohort was then stratified into exposures involving nicotine devices and nicotine liquids for the primary endpoint analysis.

**Figure 2.**
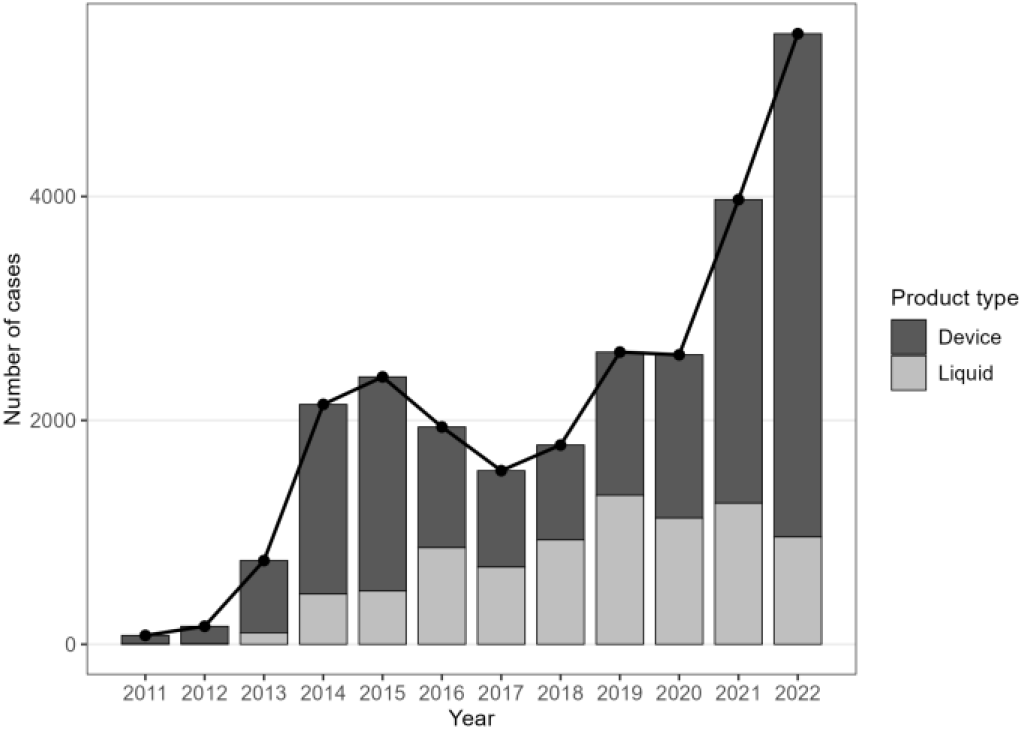
The stacked bar chart displays the annual number of cases involving nicotine liquids and nicotine devices. The overlaid black line represents the total number of combined annual exposures. The figure illustrates a shift in the primary source of exposures, with a substantial increase in device-related cases driving the overall rise in total exposures from 2018 onward.

Table 1 summarizes demographic and exposure characteristics among followed cases. The mean age was 20.6 months in the device group and 20.5 months in the liquid group (p = 0.50). Mean weight was 12.1 kg and 11.9 kg, respectively (p = 0.28). Gender distribution was similar across groups (p = 0.47). Exposure and caller site were predominantly the child’s own residence for both groups. Statistically significant differences were observed in the distributions of both caller site (p = 0.03) and management site (p < 0.001). A greater proportion of liquid exposure cases were referred to or already at a healthcare facility compared with device exposures (table 1).

**Table 1.**
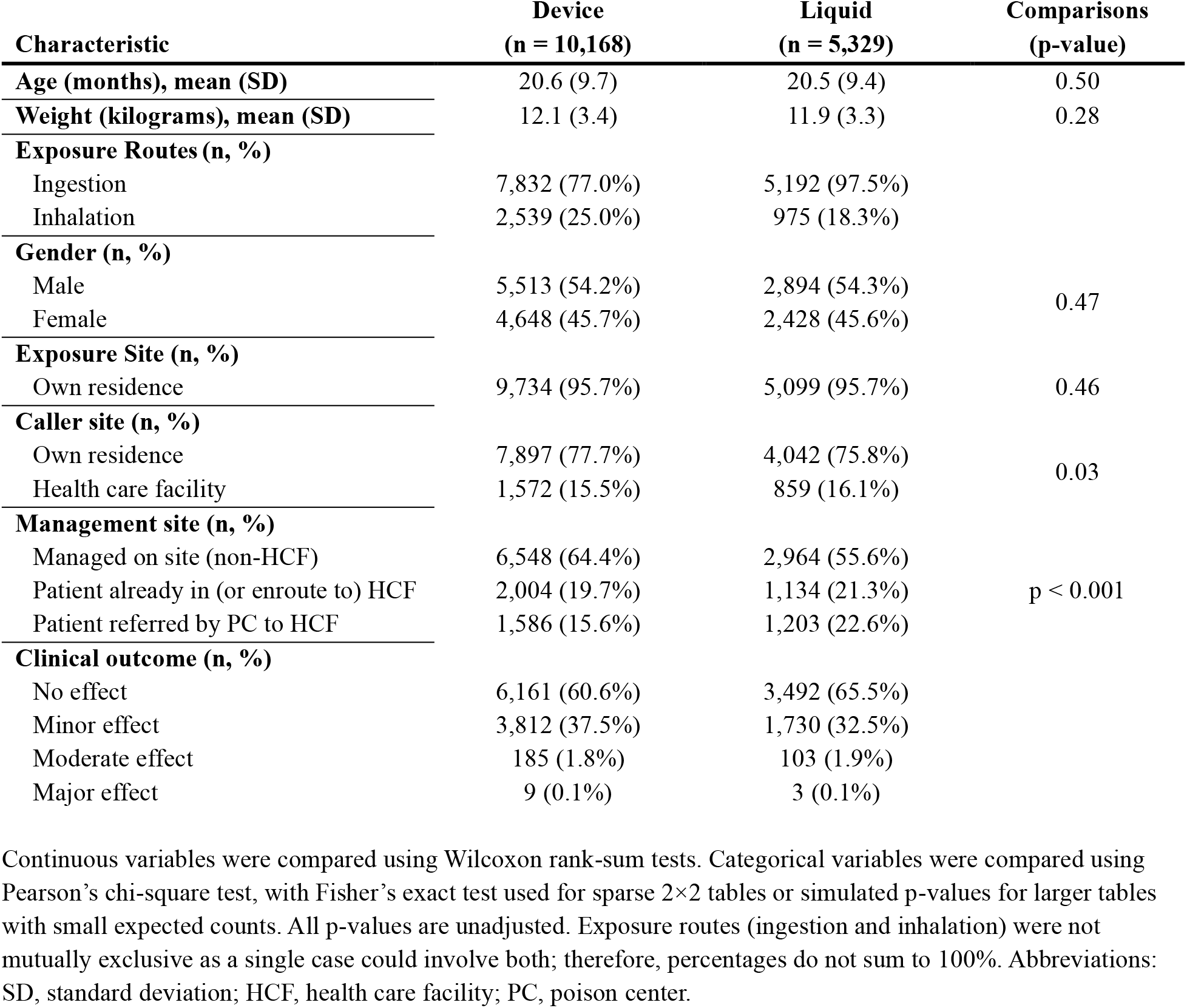
Demographic and exposure characteristics among followed pediatric nicotine exposure cases by product type.

Ingestion was the most common exposure route overall. Device exposures had more inhalation involvement (25.0%) compared with liquids (18.3%) (table 1). The five most frequently reported clinical effects were similar across groups, with cough/choke and vomiting most common (table 2). Exploratory analyses of individual clinical effects and therapies were conducted without correction for multiple comparisons. The duration of symptoms was short in most cases, with more than 95% resolving within eight hours. No substantial differences were observed in the distributions of reported clinical effects or symptom duration.

**Table 2.** Most frequently reported clinical effects and duration of symptoms among followed pediatric nicotine exposure cases by product type.

| **Clinical effect (n, %)** | **Device**<br>**(n = 10,168)** | **Liquid**<br>**(n = 5,329)** |
| --- | --- | --- |
| Cough/choke | 1,599 (15.7%) | 686 (12.9%) |
| Vomiting | 1,336 (13.1%) | 732 (13.7%) |
| Nausea | 140 (1.4%) | 63 (1.2%) |
| Drowsiness/lethargy | 114 (1.1%) | 70 (1.3%) |
| Agitation | 109 (1.1%) | 61 (1.1%) |
| **Duration (n, %)** |  |  |
| ≤ 8 hours | 2,199 (97.2%) | 1,061 (96.3%) |
| > 8 hours | 63 (2.8%) | 41 (3.7%) |

Frequencies of the five most common clinical effects and the duration of symptoms are presented as counts and column percentages n (%). For this analysis, the 'clinical effect duration' variable was dichotomized into two categories: ≤ 8 hours and > 8 hours.

The primary outcome, defined as a moderate or major clinical effect or death, occurred in 1.9% of device cases and 2.0% of liquid cases (OR 1.05; 95% CI, 0.82–1.34; p = 0.71), corresponding to an absolute risk difference of 0.1 percentage points. These findings suggest no significant difference in the risk of clinically important toxicity between product types. When the analysis was restricted to route-concordant exposures, comparing inhalation-only device exposures with ingestion-only liquid exposures, event rates remained low (1.5% vs. 2.0%; OR 1.38; 95% CI, 0.92–2.12; p = 0.12). Two deaths were observed in the cohort, one in each group. Because NPDS is a coded surveillance database, individual medical records were not available for review to further characterize these severe cases.

## DISCUSSION

The principal finding of this study is that, contrary to our hypothesis, pediatric exposures to e-cigarettes were not associated with a statistically significant difference in clinically important outcomes. This result challenges the implicit assumption that these devices are safer in an unintentional exposure. Although mortality from pediatric nicotine exposures is exceedingly rare, these cases frequently result in healthcare encounters. A national emergency department surveillance study using National Electronic Injury Surveillance System data identified 4,745 emergency department visits for e-liquid poisonings among children under five years of age from 2013 to 2017, despite few severe outcomes [13]. This pattern reflects the frequent healthcare utilization associated with pediatric nicotine exposures and suggests that some referrals may be avoidable, highlighting an opportunity to further study poison center triage. However, our findings suggest that the product’s physical form alone is not a sufficient predictor of clinical severity and therefore should not be used in isolation to guide patient management.

The route-restricted analysis produced similar findings despite excluding nearly half of the followed cohort. Although the estimated odds ratio increased modestly, confidence intervals remained wide and included the null value, suggesting that restricting the analysis to inhalation-only device exposures and ingestion-only liquid exposures did not materially alter the overall conclusions.

A likely explanation for this lack of difference in severity is that the total absorbed nicotine dose is often low and clinically comparable in typical pediatric exploratory scenarios, despite different exposure routes. A brief, unintentional puff from a modern disposable device likely delivers a small systemic nicotine dose, with measured yields ranging from 0.01 to 0.05 mg per puff across varying nicotine strengths [14]. Conversely, while a lick, sip, or taste of a highly concentrated e-liquid represents a larger potential dose, its gastrointestinal absorption is slower and incomplete due to significant first-pass metabolism. Furthermore, these liquids often induce a profound emetic effect, further limiting absorption [15]. It is plausible, therefore, that a puff with high bioavailability and a small lick with low bioavailability results in a similar, low-milligram systemic dose. In these common scenarios, both exposure types may deliver a sufficient dose to cause mild, transient symptoms such as vomiting or coughing but frequently fall below the threshold required to produce more severe toxicity.

Our data also reveal a rapid epidemiological shift in the source of pediatric nicotine exposures. While the decline in bottled e-liquid cases is consistent with the effectiveness of policies such as mandated child-resistant packaging [3], this trend may also reflect a broader market shift away from refillable systems toward sealed disposable devices. The concurrent rise in disposable device incidents suggests that the toxicological burden has not been eliminated but rather displaced to a newer product category. This evolution underscores the need for continued surveillance and research as clinical attention expands from traditional and modern nicotine products to emerging threats such as high-concentration nicotine pouches.

### Limitations

NPDS captures reported exposures rather than population incidence and therefore may be subject to reporting bias. A significant limitation is our reliance on coded surveillance data without individual chart review, which creates a risk of exposure misclassification. For example, poison center specialists may code an exposure involving a disposable device as a nicotine liquid exposure if the internal e-liquid is considered the primary toxicant. Consequently, our data may not always distinguish between direct device actuation and access to the liquid reservoir. Similarly, some cases coded as inhalation exposures may reflect legitimate mixed-route exposures or uncertainty in route classification rather than coding error alone. To address this, we performed a secondary, route-restricted analysis limited to inhalation-only device exposures and ingestion-only liquid exposures. Although this approach reduced ambiguity, it cannot fully distinguish true coding errors from legitimate overlap in exposure mechanisms and resulted in a substantially smaller analytic cohort.

Furthermore, this study’s timeframe encompasses a significant evolution in e-cigarette design. The market has shifted from early-generation, refillable consumer devices with accessible reservoirs to modern, closed-system disposable products. Our data lacked the granularity to differentiate between these device subtypes, which may have obscured temporal trends related to product-specific risks. Finally, this study was limited to commercially available nicotine products and the regulatory landscape of the United States. Consequently, findings may not be generalizable to regions with different products, patterns of use, or poison center practices.

## Conclusion

In conclusion, this study found no evidence that the shift in the consumer market from bottled nicotine e-liquids to disposable e-cigarettes has resulted in a corresponding shift in clinical severity for pediatric exposures. Our findings do not provide evidence that disposable devices are associated with lower clinical severity than bottled liquids. From a clinical toxicology standpoint, management and disposition decisions should continue to be guided by established principles, focusing on the patient’s symptoms and exposure history rather than the specific product involved.

Ultimately, product formulation appears to be a less important factor for risk stratification than the individual circumstances of the exposure.

## Data Availability

De-identified data, RStudio code markup document, and STROBE checklist are available at DOI: [10.5281/zenodo.18969736].

https://doi.org/10.5281/zenodo.18969736

## Contributors

RM contributed to study conceptualization, design, methodology, and drafting of the manuscript. RM conducted the statistical analysis. All authors contributed to manuscript editing and revision. SV provided supervision and critical review.

## Funding

The authors received no financial support for the research, authorship, or publication of this article.

## Competing interests

None declared. No funding was secured for this study.

## Prior publication

An earlier version of this data and analysis was presented as an abstract at the 2023 North American Congress of Clinical Toxicology, held in Montreal, Québec, Canada.

## Data availability statement

De-identified data, RStudio code markup document, and STROBE checklist are available at DOI: [10.5281/zenodo.18969736]. American’s Poison Centers (APC) maintains the National Poison Data System (NPDS), which houses de-identified case records of self-reported information collected from callers during exposure management and poison information calls managed by the country’s poison centers. NPDS data do not reflect the entire universe of exposures to a particular substance as additional exposures may go unreported.

## Notes

### Competing Interest Statement

The authors have declared no competing interest.

### Author Declarations

The Institutional Review Board of UT Health San Antonio waived ethical approval for this work (STUDY00002603). It was granted an exemption from formal review on the basis that it exclusively analyzed non-identifiable, de-identified data.

